# Results of intensified source finding efforts among community-acquired cases of Legionnaires’ disease – first results of the German LeTriWa study; Berlin, 2016−2019

**DOI:** 10.1101/2020.04.09.20056366

**Authors:** U. Buchholz, H. J. Jahn, B. Brodhun, A-S. Lehfeld, M. Lewandowsky, F. Reber, K. Adler, J. Bochmann, C. Förster, M. Koch, Y. Schreiner, F. Stemmler, C. Gagell, E. Harbich, W. Haas, B. Schaefer, C. Lück, on behalf of the LeTriWa study group

## Abstract

**Introduction:** Sources of infection of most cases of community-acquired Legionnaires’ disease (CALD) are unknown.

**Objective:** Identification of sources of infection of CALD.

**Setting:** Berlin; December 2016–May 2019.

**Participants:** Adult cases of CALD reported to district health authorities and consenting to the study; age and hospital matched controls.

**Main outcome measure:** Percentage of cases of CALD with identified source of infection.

**Methods:** Analysis of secondary patient samples for monoclonal antibody (MAb) type (and sequence type); questionnaire-based interviews, analysis of standard household water samples for Legionella concentration followed by MAb (and sequence) typing of *Legionella pneumophila* serogroup 1 (Lp1) isolates; among cases taking of additional water samples to identify the infectious source as appropriate; recruitment of control persons for comparison of exposure history and contents of standard household water samples. For each case an appraisal matrix was filled in to attribute any of three source types (external (non-residence) source, residential non-drinking water (RnDW) source, residential drinking water (RDW) as source) using three evidence types (microbiological results, cluster evidence, analytical-comparative evidence (using added information from controls)).

**Results:** Inclusion of 111 study cases and 202 controls. Median age of cases was 67 years (range 25– 93 years), 74 (67 %) were male. Among 65 patients with urine typable for MAb type we found a MAb 3/1-positive strain in all of them.

Compared to controls being a case was not associated with a higher Legionella concentration in standard household water samples, however, the presence of a MAb 3/1-positive strain was significantly associated (OR = 4.9, 95 % confidence interval (CI) 1.7 to 11). Thus, a source was attributed by microbiological evidence if it contained a MAb 3/1-positive strain, by cluster evidence if at least two cases were exposed to it and by analytical-comparative evidence if a case was exposed to it and the type of source was statistically significantly associated with being a case. We identified an infectious source in 53 (48 %) of 111 cases: in 16 (14 %) an external source, in 9 (8 %) a RnDW source, and in 28 (25 %) we attributed RDW. We attributed 9 cases to RnDW because cases were associated with wearing not regularly disinfected dentures (OR = 3.2, 95 % CI 1.3 to 7.8).

**Conclusion:** Using the appraisal matrix we attributed almost half of all cases of CALD to an infectious source, predominantly RDW. Risk for LD seems to be conferred primarily by the type of Legionella rather than the amount. Dentures as a new infectious source needs further, in particular, integrated microbiological, molecular and epidemiological confirmation.

## Introduction

In Germany Legionnaires’ disease (LD) is a notifiable form of pneumonia caused by Legionella bacteria. Typically, the bacterium is found in water systems or biofilm, but it must be aerosolized and inhaled, or aspirated to cause disease [1]. Transmission of Legionella is possible through a variety of sources, such as: aerosols of evaporative cooling systems [2, 3], whirlpools [4, 5] and residential drinking water (RDW; e.g. shower) [6].

Epidemiologically, three categories are distinguished: travel-associated, hospital-associated and community-acquired cases of LD (CALD), the latter comprising at least 70 % of all reported cases [7]. Sources of cases of CALD that are not part of a larger outbreak remain for the most part unexplained [8-10]. This is partly due to the fact that in the past only a genotypic match has been accepted as evidence to attribute a source, and it is often difficult to collect veritable lower respiratory samples (for culture and PCR) in addition to water samples from all potential infectious sources.

The objective of this study is to find out in how many cases of CALD we can identify an evidence-based source of infection.

## Methods

### Study partners

The LeTriWa study is a joint project of the Robert Koch Institute (RKI), the German Environment Agency (GEA) and the Legionella Consulting Laboratory (LCL) working in close cooperation with the Berlin State Department for Health and Social Affairs (LAGeSo), the 12 Berlin district health authorities (DHA) and Berlin hospitals. Apart from the substudy presented here the LeTriWa study comprises other substudies that will be presented elsewhere.

### Definitions

#### Definition of a study case

A study case was defined as a reported case of LD and laboratory evidence of Legionella infection fulfilling the following characteristics:

- Having been reported to a Berlin DHA between 01 December 2016 and 31 May 2019 18 years or older
- Sufficient information to classify the case as travel-associated, hospital-associated or CALD (including nursing home-associated)
- In the context of the LeTriWa study: Overnight stay at home for at least four nights (arbitrarily set) and no hospitalization (defined as at least one overnight stay in the hospital) in the two weeks before onset of illness
- Ability to adequately communicate with study staff (no dementia, adequate command of the German language)
- Written consent for study participation

### Type of water sample

The German technical document DIN EN ISO 19458 defines different water sampling types from DW plumbing systems. „First flush” sample is defined as the DW sample collected immediately after opening the faucet (“as it is consumed”) [11]. It is particularly intended to help investigating the water for source finding in the case of an illness. A water sample after flushing of one liter (flushed sample) has the goal to sample water from the riser, i.e. before the distal end.

All water samples were analyzed for the concentration of Legionella at the laboratory of the GEA according to DIN EN ISO 11731. *Legionella pneumophila* serogroup 1 (Lp1) and selected serogroup (sg) 2–15 as well as non-pneumophila isolates were sent to the LCL for subtyping. LCL picked up to 20 colonies per water system and analysed for serogroup, MAb subtype [12] and sequence type (ST) [14, 15].

### External source

A source of water that is located outside the residence of an individual, e.g. water from the faucet of a sink at the case-patient’s working place, or the shower in a swimming pool visited.

### Residential non-drinking-water source

A residential non-drinking water (RnDW) source is a source, which is located within the residence of the case-patient and has no direct connection to the DW supply, but – for example – may have been sitting for a long time in a container, e.g. water of a watering can, a water filter or a humidifier.

### MAb type and MAb subtypes

Lp1 bacteria can be grouped into MAb groups according to the Dresden panel. An important group are the MAb 3/1-positive strains that are believed to be particularly virulent. Among MAb 3/1-positive bacteria four MAb-subtypes are distinguished: “Knoxville”, “Philadelphia”, “Benidorm” and “France/Allentown”.

### Course of the study and laboratory methods

#### Clinical microbiological diagnosis

Cases of LD were diagnosed in the hospitals by using urinary antigen detection and if positive were reported by the laboratory to the responsible DHA. Additional (second set) urine and lower respiratory samples were requested to be sent to the LCL. The urines were concentrated 10-fold and heated for 15 min at 95 °C. An in-house ELISA tested for the presence of MAb 3/1-positivity (according to the Dresden panel) [12] and – if positive – for the MAb subtype [13].

Respiratory samples were tested by culture and PCR for *Lp1*. In samples with medium or high DNA content direct sequence-based typing was conducted.

### Search for infectious sources

First DHA and (after giving consent) also RKI contacted the patients. After having received written consent to the study we conducted a detailed questionnaire inquiring about exposures in the 14 days before symptom onset (period of infection; see supporting materials). In addition, we took standard household water samples including a first flush, biofilm and flushed sample from the faucet from the sink, as well as a first flush and biofilm sample from the shower. If applicable, further samples were taken from additional DW or non-DW sources or external sources.

In order to allow the DHA to gauge the situation and to take measures if appropriate, a complete expert risk assessment of the DW plumbing system in the residence of the case was carried out wherever possible [16].

### Control group

For each case we recruited two controls matched by hospital and age group (< 50 years, 50–74 years, > 74 years), who were admitted for a reason other than pneumonia. For control persons the same questionnaire was used as for study cases. In the household of controls we took only standard household water samples.

### Appraisal of results

We have conceptually organized the attribution of potential sources into a matrix of source types and evidence types (Table 1). Potential sources of infection include (1) “external source”, (2) “residential non-DW source”, and (3) “residential DW source”. Evidence types include (1) microbiological evidence, (2) cluster evidence, and (3) analytical-comparative evidence.

**Table 1:** Matrix to classify source types and evidence types; RnDW = residential non-drinking water, RDW = residential drinking water.

| <u>source type</u> | <u>evidence type</u> |  |  |
| --- | --- | --- | --- |
|  | microbiological evidence | case is part of a cluster | analytical-comparative evidence |
| external source |  |  |  |
| RnDW source |  |  |  |
| RDW source |  |  |  |

The evidence types are explained as follows:

#### Microbiological evidence

In a given case, results from the patient, from the sample of the potential source and – if available – the comparison of the two were evaluated (Table 2). Previously it was shown that the majority of isolates in human infections in Germany were MAb 3/1-positive, while most environmental isolates taken from routine water samples in Germany were not [17]. As also all patients’ MAb types in typeable urine samples were MAb 3/1-positive and being a case was significantly associated with the presence of a MAb 3/1-positive standard household water samples (see Results), we attributed a potential source of infection to a case when we found a MAb 3/1-positive strain in a presumptive source, even when we were lacking information on the patient’s MAb type (Table 2; category 2). Should we find a different MAb type, subtype, or sequence type in the patient compared to the infection source this source was not attributed to the case.

**Table 2:** Microbiological evidence by category of identified strains among cases and potential environmental infectious sources; increasing degree of evidence from category. Cat. = Category, Lp1 = L. pneumophila serogroup 1, MAb = monoclonal antibody, ST = sequence type.

| Cat. | Case strain | Environmental source strain | Source attributed to case | Nr of cases by category |
| --- | --- | --- | --- | --- |
| 1 | irrelevant | no isolate that is MAb 3/1+ or ST of case and source incongruent or MAb type or subtype of case and source incongruent | No | 72 |
| 2 | Lp1, MAb type and sequence type unknown | at least one isolate is MAb 3/1+ | Yes | 12 |
| 3 | Lp1, MAb 3/1+, MAb subtype unknown, ST unknown | same MAb type as case | Yes | 5 |
| 4 | Lp1, MAb 3/1+, MAb subtype known, ST unknown | same MAb subtype as case | Yes | 15 |
| 5 | ST known | same ST as case | Yes | 7 |

#### Cluster evidence

A case was part of a cluster, defined as same exposure as at least one other case to a potentially infectious source (within two years of dates of symptom onset). The other case(s) belonging to the cluster may be other reported, non-study cases of LD.

#### Analytical-comparative evidence

As a third way to assign a source of infection to a case we compared the frequency of exposure to a possible source of infection in cases with that of controls. The type of exposure is extracted for the most part from the questionnaires, but may also be exposure to – for example – MAb 3/1-positive water sampled from the faucet in the bathroom. If an exposure variable is then identified as statistically significant, the corresponding source of infection is attributed to all the cases where this variable applies.

In the case when more than one possible infectious source could be attributed to a given case we assigned a source individually based on the available microbiological, cluster and/or analytical-comparative evidence.

#### Non-Responder-Analysis

We compared age, gender and place of residence of CALD who could have participated in the study but refused to participate with that of study cases (who by definition agreed).

#### Data handling and statistical analysis

We entered data in Microsoft Excel (Microsoft Office Professional Plus 2010, Redmond, WA, USA). Data were analysed in Microsoft Excel or in Stata, version 15 (Stata Corporation, College Station, TX, USA). Statistical analysis was largely descriptive. Missing data were not imputed. To evaluate the analytical-comparative evidence we carried out bivariate analyses of relevant variables computing odds ratios (OR) for disease, complemented by stratified analyses as appropriate.

### Quality assurance

RKI staff was certified to take samples according to DIN EN ISO 19458 [11]. The GEA laboratory is accredited by the German Accreditation Body DAkkS for testing DW in accordance with Drinking Water Ordinance (2001) [18]. Except for external sources employees of the GEA laboratory were blinded with regard to the case or control status of water samples. Taking of water samples from external sources and the in-building risk assessments were carried out by an accredited hygiene engineer commissioned by GEA. He and his employees were not informed whether the DW plumbing system of a building to be investigated belonged to a case or a control person.

### Ethics approval

Ethics approval for the study was granted by the Ethics Committee of the Charité (EA1/303/15). In addition, a data protection concept was drawn up with the data protection officer of the RKI and then submitted to and approved by the Federal Commissioner for Data Protection and Freedom of Information.

### Registration of the study in the German Registry of Clinical Studies

The study is registered in the German Registry of Clinical Studies with the identifier DRKS00009831 (https://www.drks.de/drks_web/navigate.do?navigationId=trial.HTML&TRIAL_ID=DRKS00009831).

## Results

Between 01 December 2016 and 31 May 2019 (30 months) the Berlin 12 DHA reported 330 laboratory confirmed cases of LD to the RKI. After subtraction of the cases not meeting the inclusion criteria 190 cases remained that were eligible for inclusion in the study. Of these, 79 (42 %) refused to participate and 111 (58 %) agreed (study cases) (Figure 1). Comparison of the non-responders (n = 79) with the 111 study case-patients showed no significant differences in age, gender and district. For comparison we recruited 202 controls. Cases and controls did not differ in respect to age group, gender and district.

**Figure 1:**
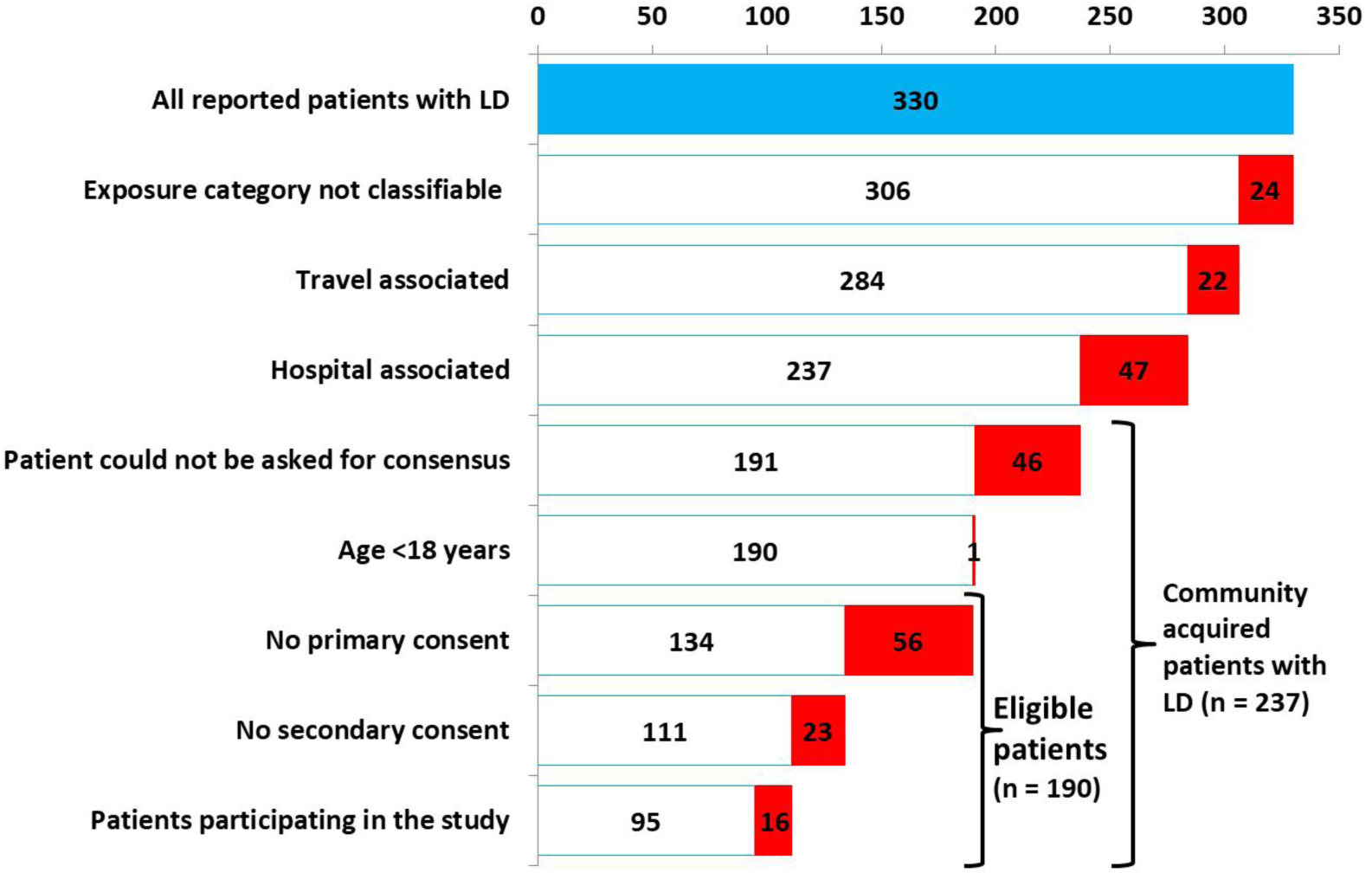
Number of reported patients, eligible patients and patients participating in the study; LD = Legionnaires’ disease. In red: number of patients where the respective category applies; Berlin, 2016–2019.

Study cases (n = 111) who agreed to participate in the study had a median age of 67 years (range 25– 93 years), 74 (67 %) were male. Thirty (27 %) had no previous underlying/predisposing disease, 63 (57 %) had a moderately predisposing underlying disease (e.g. preconditions of the lung or heart) and 18 (16 %) had a strongly predisposing underlying disease (immune deficiency). Fifty-three (48 %) study cases smoked. In 67 (60 %) of the 111 study cases lower respiratory samples were available, these were only in 19 (28 %) tracheal secretion or from bronchoalveolar lavage (BAL). We could identify a sequence type in 16 (24 %) cases with lower respiratory samples, these were 9 times ST182 (“Berlin clone”) and 7 times other STs.

LCL received urine samples from 100 (90 %) of the 111 study cases. In 85 (85 %) of the 100 study cases with urine samples the Legionella infection could be confirmed by urinary antigen test. In 65 (76 %) of the 85 study cases with confirmed Legionella infection in the LCL, we could confirm MAb 3/1-reactivity. In 40 of these the MAb subtype could be defined and was always “Knoxville”.

We took standard household water samples in all study cases, household non-RDW samples in 44 (40 %) of all 111 study cases and samples from external infection sources in 43 (39 %) of 111 study cases. Expert risk assessments of the DW plumbing system was done in 41 (37 %) of all 111 study cases.

### Microbiological evidence

Based on microbiological evidence we attributed 39 times an infectious source to a case (Table 2, Table 3). Among the 39 (35 % of 111) cases with attributable source through microbiological evidence we attributed 12 (31 %) solely based on the finding of a MAb 3/1-positive strain in a water source having exposed the patient (Table 2). Further 5, 15 and 5 cases had an attributable source because MAb type (MAb 3/1), MAb subtype or ST, respectively, matched between patient and water sample. In three cases the MAb subtype had matched, however, the ST did not, so the respective water source was not attributed on microbiological grounds.

**Table 3:**
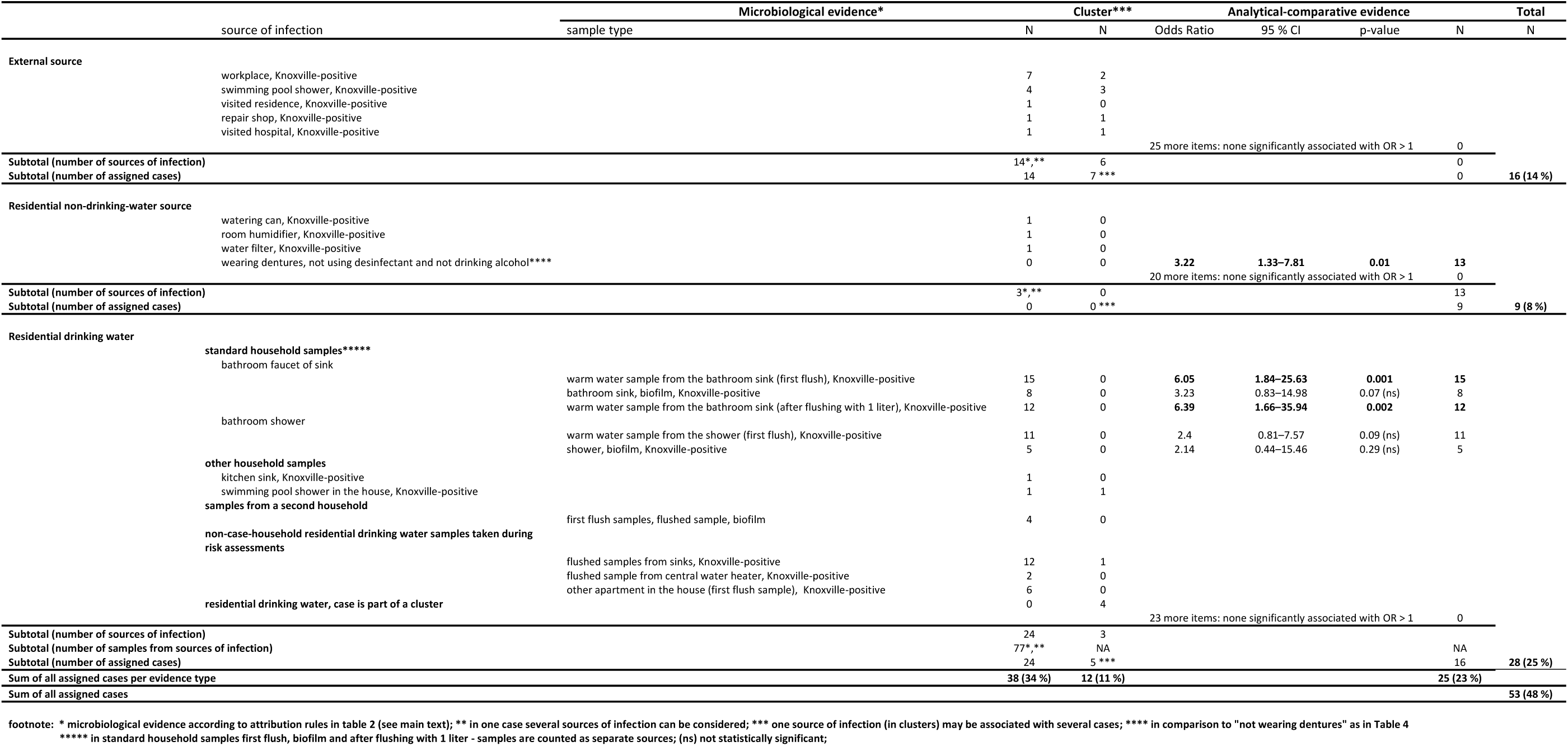
Infection sources of 111 study cases broken down by category (external, residential non-drinking water, residential drinking water) and evidence type (microbiological, cluster, analytical-comparative); Note that regarding residential drinking water standard household water samples were taken in every case, while “other household samples” and “samples during risk assessment” were taken only in 44 and 41 cases, respectively; Berlin, 2016–2019.

### Cluster evidence

We identified 12 patients who had an exposure to an infectious source that was shared by at least one other person. This led (in synopsis with the results of the investigations of the 126 (237–111) non-study cases) to the detection of 8 clusters: 6 clusters (63 %) among cases with evidence for infection through an external source, no cluster with evidence for infection through a RnDW source and three clusters (33 %) with evidence for infection through RDW. Because one cluster was technically an external source for one patient and RDW for another, the total number of clusters was not 9, but 8. The size of the clusters was 6 times 2 patients and 2 times 4 patients.

### Analytical-comparative evidence

Based on results from the questionnaires we identified one source (not disinfected dentures) that was significantly associated with case status (see below under the section “Residential, non-drinking water sources as sources of infection of CALD”).

### External water sources as sources of infection of CALD

Microbiological evidence: Overall we took samples from 58 sources among 43 (39 %) of all 111 study cases, thus we sampled 0.52 external sources per study case (58/111). In 14 (13 %) of the 111 study cases, we found a Lp1 strain that was MAb 3/1-positive in one of the relevant external water sources (microbiological evidence; Table 3), in four cases a possible infecting external source was not attributed because the ST of the water source (ST182) did not match that of the patient (ST was undefined, but 182 was excluded). All 18 infection sources of these 18 cases were DW sources and none were non-DW sources.

Exposure to MAb 3/1-positive sources of infection occurred in seven (50 %) of the 14 cases at the workplace (including one hospital), in four cases (29 %) in a shower of a swimming pool, in one case (7 %) in a visited residence, in one case (7 %) in a repair shop and in one case (7 %) in a visited hospital. A MAb 3/1-positive strain was found in five (36 %) of the 14 cases in shower water (of visited swimming pools or at the workplace), in eight (57 %) in faucet water, in one (7 %) in faucet water and at the same time in shower water.

#### Cluster evidence

Four clusters included a total of five study cases for whom also microbiological evidence existed (resulting in five (36 %) of 14 cases with microbiological evidence belonging to a cluster); the other cases contributing to these four clusters were non-study cases. A further two cases belonged each to an external source cluster where the cluster definition was met because non-study cases contributed. In total, seven (of 16 (44 %) study cases were assigned to an external source because of cluster evidence (Table 3).

#### Analytical-comparative evidence

being a study case was not statistically significantly associated with any of 25 items from the questionnaire representing potential external sources of infection.

### Residential non-drinking water sources as sources of infection of CALD

Microbiological evidence: Overall we took samples from 57 sources, on average from 0.51 RnDW sources per study case. We found Legionella bacteria in eight (14 %) of the 57 possible sources. In three cases we identified a MAb 3/1-positive RnDW source, a humidifier, a watering can and a water filter (Table 3), however, they were not attributed as infecting source because also the RDW contained MAb 3/1-positive strains; therefore the RDW was attributed in these cases. We tested nine dentures or the containers where these were kept, but could not identify Legionella in any. None of these nine belonged to the nine cases that were assigned to a RnDW source (dentures) based on analytical-comparative evidence (see below).

#### Cluster evidence

No RnDW infection source was associated with a cluster.

Analytical-comparative evidence: In bivariate analysis the OR for wearing dentures was 1.93 (95 % CI = 1.2 to 3.2, p-value = 0.009). Controlling for age group, even for five year age groups, did not alter the effect. In stratified analyses, we found that the effect of wearing dentures was modified by two other variables: (1) laying dentures in a disinfectant solution, and (2) drinking alcohol, i.e. not being abstinent from alcohol). Thus, being a study case was significantly associated with the characteristic “wearing dentures and not disinfecting them and not being abstinent from alcohol” (OR = 3.2, 95 % CI = 1.3 to 7.8, p-value = 0.01). This characteristic applied to 13 cases (Table 4). Twenty further additional variables that captured possible exposures to non-DW sources were not associated with being a study case.

**Table 4:** Association of wearing dentures, with/without cleaning them in disinfectant, being abstinent from alcohol (or not), and Legionnaires’ disease; Berlin, 2016–2019.

| characteristic | wearing dentures | using disinfectant | alcohol consumption | Odds Ratio (95 % confidence interval) | p-value |
| --- | --- | --- | --- | --- | --- |
| not wearing dentures (reference) | no | irrelevant | irrelevant | 1 |  |
| wearing dentures and cleaning them in disinfectant, drinks alcohol | yes | yes | yes | 0.6 (0.23–1.54) | 0.28 |
| wearing dentures and cleaning them in disinfectant, abstinent from alcohol | yes | yes | no | 2.26 (0.9–5.63) | 0.08 |
| wearing dentures and not cleaning them in disinfectant, drinks alcohol | yes | no | yes | 1.36 (0.61–3.05) | 0.45 |
| wearing dentures and not cleaning them in disinfectant, abstinent from alcohol | yes | no | no | 3.22 (1.33–7.81) | **0.01** |

### Residential drinking water sources as sources of infection of CALD

Microbiological evidence: We identified MAb 3/1-positive samples in different frequencies from all types of standard household water samples (Table 3). In addition, there were individual MAb 3/1-positive samples in other locations, such as from a swimming pool shower of a residence or from the faucet of a kitchen sink. Water samples taken during risk assessment of the DW plumbing system have verified evidence of standard household samples in 11 cases and led to further RDW attributions in two study cases. In no case did the patient’s ST contradict the ST of the DW strain that led to attribution of DW to the cases. In sum, we had microbiological evidence to attribute RDW as infecting source for 24 study cases.

#### Cluster evidence

One study case belonged to a cluster with additional microbiological evidence (Table 3). Four more cases had cluster evidence without microbiological confirmation: two cases each had been exposed to the RDW of the same DW plumbing system, respectively. Thus, a total of five cases were assigned RDW as source based on cluster evidence.

#### Analytical-comparative evidence

Comparing study cases with the control group we found that cases were significantly more frequently exposed to MAb 3/1-positive standard household water samples (Table 5, top). Cases were not significantly associated with exposure to any Legionella bacteria and Lp, but were significantly associated with the variable “Lp1 in any standard household sample” (Table 5, top). The OR gathered around 1 (the null hypothesis) when the analysis was restricted to cases and controls without MAb 3/1-positive standard household samples (Table 5, bottom). Overall, there was double (microbiological and analytical-comparative) evidence in 16 cases (14 %). Cases were not more likely to have higher concentrations of Legionella in standard household water samples. Twenty-three further additional variables that captured possible exposures to RDW as a source were not associated with being a study case.

**Table 5:** Association of Legionnaires’ disease and exposure to different types of contamination with Legionella bacteria in standard household water samples; CI = confidence interval, sg = serogroup, MAb = monoclonal antibody; Berlin, 2016–2019.

| Exposure | Cases |  |  | Controls |  |  | Odds | 95 % CI | p-value |
| --- | --- | --- | --- | --- | --- | --- | --- | --- | --- |
|  | Total | Exposed | % | Total | Exposed | % | Ratio |  |  |
| All study cases and controls samples |  |  |  |  |  |  |  |  |  |
| any Legionella | 111 | 60 | 54 | 172 | 100 | 58 | 0.85 | (0.51–1.4) | 0.54 |
| *L. pneumophila* | 111 | 45 | 41 | 172 | 58 | 34 | 1.34 | (0.79–2.3) | 0.26 |
| *L. pneumophila* sg 1 | 111 | 41 | 37 | 172 | 44 | 26 | 1.70 | (0.98–3.0) | 0.05 |
| *L. pneumophila* sg 1, MAb 3/1+ | 111 | 20 | 18 | 161 | 8 | 5 | 4.97 | (1.68–11) | 0.001 |
| Study cases and controls excluding those with at least one MAb 3/1-positive sample among standard household samples |  |  |  |  |  |  |  |  |  |
| any Legionella | 91 | 40 | 44 | 153 | 82 | 54 | 0.68 | (0.39–1.2) | 0.19 |
| *L. pneumophila* | 91 | 25 | 27 | 153 | 50 | 33 | 0.78 | (0.42–1.4) | 0.47 |
| *L. pneumophila* sg 1 | 91 | 21 | 23 | 153 | 36 | 24 | 0.97 | (0.50–1.9) | 1 |

### Discrepant pieces of evidence

By virtue of the concept of the matrix more than one infection source could have been identified for an individual patient. We found the following conflicts:

1. In two study cases we identified a RnDW source as well as a RDW source. In those cases we decided that the RDW source was more likely to have caused the cases of LD.
2. In two study cases we had analytical-comparative evidence for dentures as infecting source, at the same time there was microbiological evidence for RDW. In these we gave preference to RDW as the more likely source.
3. In one case we had analytical-comparative evidence for dentures as infecting source, at the same time the study case was part of a RDW-cluster. The case was therefore attributed to RDW.
4. In one study case we had evidence for three possible sources: (a) analytical-comparative evidence for dentures as infecting source, (b) microbiological evidence for RDW, and (c) microbiological evidence for RnDW. Again we gave preference to the RDW.

In sum, 38 (34 %) of the 111 study cases could be attributed by microbiological evidence, 12 (11 %) by cluster evidence and 25 (23 %) by analytical-comparative evidence (Table 3). Broken down by source type, 16 (14 %) of 111 cases could be assigned to an external DW source, nine (8 %) to a RnDW source and 28 (25 %) to a RDW source for a total of 53 (48 %) of 111 study cases with an identified infection source, the breakdown by source type and (single or combined) evidence type of all study cases is shown in Figure 2.

**Figure 2:**
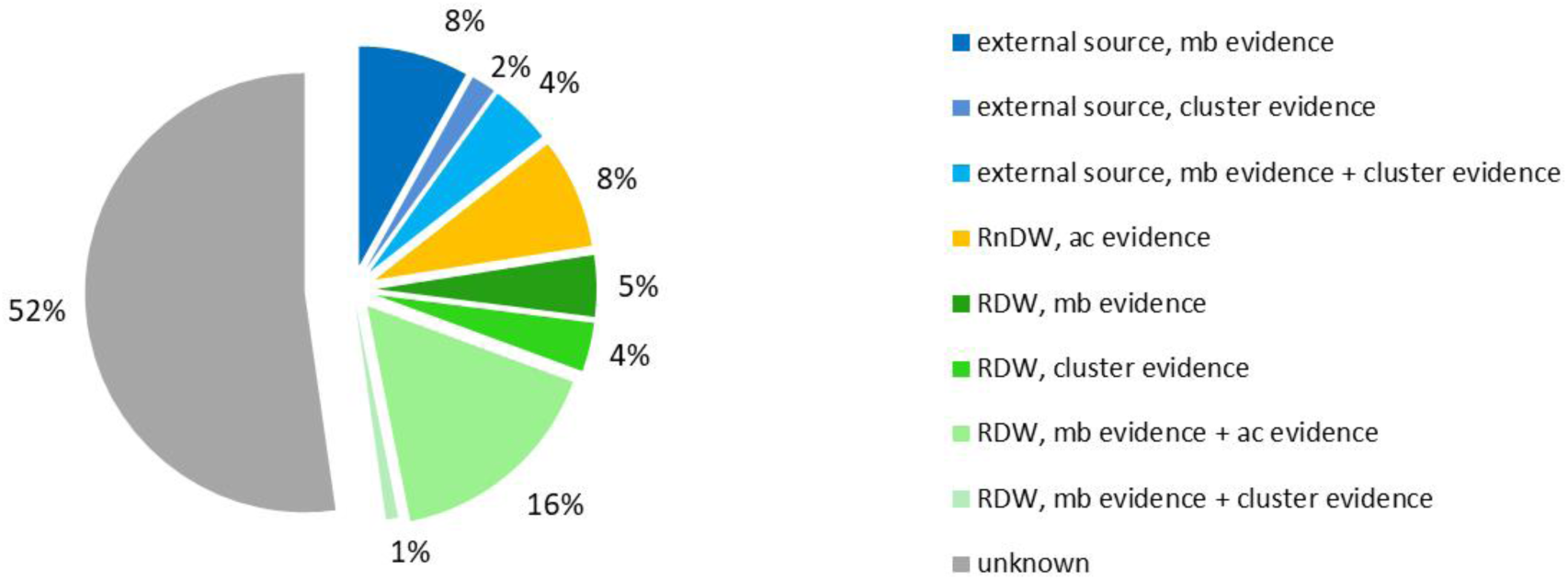
Infection sources by category (external (blue), residential non-drinking water (RnDW, orange), residential drinking water (RDW, green)) and evidence types (microbiological (mb), cluster, analytical-comparative (ac)); Berlin, 2016–2019 (N = 111).

## Discussion

Based on our thorough investigations and conceptual approach we have attributed approximately half of all Berlin cases of CALD to a likely source. Of the cases with an attributed source more than half has probably been infected through RDW. The most outstanding characteristic of RDW leading to infection was MAb 3/1-positivity, whereas the degree of Legionella concentration in the standard household water samples was not relevant. Cases not attributed to RDW were attributed to either varying external sources or to wearing dentures if these were not disinfected.

We have devised a novel, comprehensive concept where we have distinguished three source types (external, RnDW, RDW) which we attributed to a given case using at least one of three evidence types (microbiologically, cluster, analytical-comparative). The source categories have been chosen with a specific reasoning. It is important to understand what proportion of cases is infected away from home versus at home, and among the latter which proportion is infected due to the direct influence of DW. With respect to the type of evidence, traditionally only microbiological evidence (a genotypic match) was used to attribute a source to a single case. Similar to den Boer et al. we have also included the occurrence of belonging to a cluster as evidence, i.e. when two cases were exposed to the same source (within two years of dates of symptom onset) [9]. Because we had collected also data from a control group we added a third evidence type, analytical-comparative evidence. These three evidence types support each other in many study cases. This led to an attribution rate of approximately 50 % which we believe is unprecedented in the literature.

Clearly, the most abundant evidence exists for RDW. A priori we were unsure about the significance of finding “any contamination” with Legionella bacteria (irrespective of the degree of contamination), a certain degree of contamination or the presence of specific strains (Lp, Lp1 or specific MAb types or subtypes) in water samples for the occurrence of a case. Because we collected the same standard household water samples in case households and control households we were able to analyse if being a case was associated with these Legionella parameters. We did see a difference between cases and controls for “any Legionella”, Lp and Lp1: The OR rises with increasing specificity of the parameter (“any Legionella” < Lp < Lp1) and is even statistically significant for Lp1 (Table 5, top). However, the difference is convincing only for Lp1 when the samples were MAb 3/1-positive, suggesting that the differences seen in the other parameters are confounded by MAb 3/1-positivity. Indeed, when we analyzed the parameters “any Legionella”, Lp and Lp1 for those cases and controls without any MAb 3/1-positive standard household water sample the OR converged around 1. Moreover, being a study case was not associated with higher Legionella concentrations among standard household water samples. In sum and in line with Harrison [19], this suggests that a risk exists in a given water source only if it contains a MAb 3/1-positive strain. Therefore we felt it was also justifiable to attribute a water source, no matter if it was an external source, RnDW-source or RDW-source, to a case if it contained a MAb 3/1-positive strain.

RDW is the only source type where evidence comes from all three evidence types. Among study cases we identified a MAb 3/1-positive strain significantly more frequently in two of the five standard household samples and this applied to 16 cases. The added analytical-comparative evidence strengthens substantially the evidence for the cases attributed to RDW.

The fact that water samples taken from the bathroom faucet were implicated on microbiological and analytical-comparative grounds does of course not prove that the faucet water indeed infected the patient. It only shows that the water in the DW system contained virulent Legionella strains, and did so more frequently than among controls. It is possible that transmission took place from breathing in or aspiration of kitchen or shower water.

We have attributed external sources to study cases in 14 %, in particular showers in swimming pools, and also water sources at the workplace, including faucets of hand washing sinks. It is noteworthy that seven (44 %) of the 16 cases attributed to an external source were part of a cluster, likely because these were often water sources where many people are generally exposed to, e.g. showers of a swimming pool. In a local context it may thus be helpful to investigate potential external sources as other cases may also be associated with it. In addition we wish to point out that we have – after a thorough literature search – included a large number of potentially infecting external sources in the questionnaire. However, none of them has been statistically significant.

We have tested microbiologically (on average) a RnDW source in about one of three cases and were thus relatively conservative with taking this type of samples. Although we found Legionella in several sources (including a MAb 3/1-positive strain in two), ultimately we attributed (based on microbiological or cluster evidence) none of the sources to a case. Using the analytical-comparative approach we found a robust association of being a study case with wearing dentures, particularly when these were not disinfected through a disinfecting solution and if the patient stated not to drink alcohol. Although these results are striking they still need microbiological grounding.

When a genotypic match is required for microbiological evidence typeable Legionella isolates are necessary both from patients and the suspected environmental sample which leads to a paucity of microbiologically proven sporadic cases [8-10]. We accepted in this study identification of a MAb 3/1-positive Legionella strain in a water source as microbiological evidence because its frequency is significantly higher in case households compared to control households. This led to a total of 34 % of study cases with attributed source based on microbiological grounds (Table 3).

As mentioned above we defined evidence of a cluster only in those cases where there was a common exposure to the same potentially infectious source. Den Boer called this type of cluster “location cluster” [9]. Because of methodological differences our proportion of 11 % (12/111) of cluster cases among all study cases cannot be compared directly to the proportion provided by den Boer who reported 266 (13 %) cluster cases among 1991 non-nosocomial cases of CALD. Nevertheless proportions are in the same range.

With our third evidence criterion, analytical-comparative evidence, we attributed eight study cases to the wearing of dentures. Through detailed data analysis we found that “wearing dentures” as a risk factor is not a random finding, a proxy for something else or confounded by age. Only those persons who do not disinfect their dentures seem to be at risk. This suggests that Legionellae hide in the surfaces of the dentures from where they might be inhaled. Interestingly, also the – initially – surprising finding of a protective effect of alcohol consumption might be explained if alcohol disinfects Legionella bacteria sitting on dentures. Although we have gathered interesting epidemiological evidence so far, of course, this needs to be confirmed by microbiological evidence, for example proof of Legionella in dentures.

A certain limitation of our attribution concept is that it is in principle possible that for one given case two source types are attributed and “compete” with each other. However, we were able to resolve the few instances where these conflicts occurred. On the other hand, using three evidence types opens the possibility to strengthen the support to a given infection source.

Another limitation is that our study was confined to the city of Berlin. Since both Legionella ecology and patient strains may differ from one geographic region to the other [20, 21], and the Berlin clone ST182 seems to be rather unique for this area it is unknown to what extent our findings can be generalized to other regions in Germany or other countries. However, we believe that many results, such as the association of disease with the presence of virulent (Mab 3/1-positive) strains in drinking water, but not Legionella concentration per se, are generalizable.

Overall, using our concept, and in this setting in the city of Berlin we could attribute half of cases of CALD to a source. It has led to a new hypothesis of a source (wearing dentures) and it has shown clearly the importance of DW as a source for LD. About half of all study cases remain unexplained. Judging from the findings of household water samples it is not the contamination with or the amount of Legionella bacteria that puts individuals at risk but rather the type of Legionella bacteria present, in Berlin the MAb 3/1-positive “Knoxville” strain. Further research and/or analyses are needed to underpin the role of wearing dentures for acquiring LD and to understand which factors contribute to the contamination of DW with pathogenic Legionellae in the household.

## Data Availability

The data availability is in accordance with the German data protection law and is only available to the LeTriWa study team specified in the approved data protection concept.

## Supporting information

Appendix 1: List of questions asked in the questionnaire to cases and controls.

Appendix 2: Microbiological results of all patients as well as the result of the “best” water sample taken. The “best” water sample is the one taken from a source with Legionella strains most likely having caused the infection.

## Collaborators

The LeTriWa study group: U. Buchholz1*, H. J. Jahn^1^*, B. Brodhun^1^, A-S. Lehfeld^1^, M. Lewandowsky^1^, F. Reber^1^, K. Adler^2^, J. Bochmann^2^, C. Förster^2^, M. Koch^2^, Y. Schreiner^2^, F. Stemmler^2^, C. Gagell^3^, E. Harbich^3^, S. Bärwolff^4^, A. Beyer^5^, U. Geuß-Fosu^6^, M. Hänel^7^, P. Larscheid^8^, L. Murajda^9^, K. Morawski^10^, U. Peters^11^, R. Pitzing^12^, A. von Welczeck^13^, G. Widders^14^, N. Wischnewski^15^, I. Abdelgawad^14^, A. Hinzmann^11^, D. Aguiar Pineda^13^, B. Schilling^4^, S. Schmidt^5^, J. Schumacher^8^, I. Zuschneid^15^, I. Atmowihardjo^16^, K. Arastéh^17^, S. Behrens^17^, P. Creutz^18^, J. Elias^16^, M. Gregor^17^, S. Kahl^16^, H. Kahnert^17^, V. Kimmel^17^, J. Lehmke^17^, P. Migaud^17^, A. Mikolajewska^18^, V. Moos^18^, M.-B. Naumann^17^, W. Pankow^17^, H. Scherübl^17^, B. Schmidt^16^, T. Schneider^18^, H. Stocker^17^, N. Suttorp^18^, D. Thiemig^17^; senior authors: W. Haas^1^**, B. Schaefer^2^**, C. Lück^3^**

## Acknowledgement

We are grateful to the support and advice given by Matthias an der Heiden, Jörg Lekschas, Helmut Fouquet, Katharina Schmitt, Antonia Mehlitz, Marie Reupke, Karina Preußel (all working at the RKI); the district health authorities in Melle (Lower Saxony), Munich (Bavaria), Greifswald (Mecklenburg Western Pomerania), Oranienburg and Koenigs-Wusterhausen (Brandenburg); as well as the experts in a scientific preparatory meeting (Petra Brandsema, Gavin Dabrera, Sjoerd M. Euser, Martin Exner, Wolfgang Hentschel, Klaus Heuner, Thomas Kistemann, John Lee, Birgit Mendel, Gunnar Meyer, Marius Hausner, Jürgen Thelen, Jost Wingender); Dirk Werber, Daniel Sagebiel and Claudia Simon from the Berlin State Department for Health and Social Affairs (LAGeSo); the cooperating hospitals and all district health authorities in Berlin. We also thank Katie Jacques, Anna Nolden and Corinna Fruth for helping in the field work of the study.

## Dissemination declaration

During the ongoing study: (1) The cases are provided with the number of the health authority where they can obtain information of the results of the drinking water tests of their household. (2) The property management of the cases receives the results of the systemic drinking water tests and of the technical assessment of the drinking water installation. (3) The controls and their property managers receive the results of the drinking water examinations and the technical assessment of the drinking water system from the LeTriWa study team.

After completing the data collection, published (aggregated) results of the study will be sent to study participants.

## Patient and Public Involvement statement

It was not appropriate or possible to involve patients or the public in the design, or conduct, or reporting, or dissemination plans of our research.

